# Global Learning Opportunities Within Social Innovation in Health (GLOWS): Modified Delphi Process to Identify and Pilot Core Competencies for Learning

**DOI:** 10.1101/2025.02.27.25323020

**Authors:** Emily Wallace, Yusha Tao, Ogechukwu B. Aribodor, Zixuan Zhu, Angelica Borbón, Beatrice Halpaap, Bertha M. Chakhame, Eunice C. Jacob, Fatema Ahmed, Joel Msafiri Francis, Komang G. Septiawan, Kovey Mawuli, Linet Mutisya, Marlita Putri Ekasari, Nwadiuto Okwuniru Azugo, Tina Fourie, Adriana S. Ruiz, Jackeline Alger, Abigail Ruth Mier, Weiming Tang, Gloria Aidoo-Frimpong, Jackie Nono, Jesson James A. Montealto, Obidimma Ezezika, Per Kåks, Wenjie Shan, Jana Deborah Mier-Alpano, Gifty Marley, Elizabeth Chen, Joseph D. Tucker

**Affiliations:** London School of Hygiene and Tropical Medicine, London, United Kingdom; The Spark Innovation Programme, Health Service Executive, Ireland; University of North Carolina Project-China, Guangzhou, China; Dermatology Hospital of South Medical University, Guangzhou, China; Department of Zoology, Nnamdi Azikiwe University, Nigeria; Social Innovation in Health Initiative (SIHI) Nigeria Hub, Nigeria; Innovation in Public Health, Department of Research in Public Health, National Institute of Health, Bogotá, Colombia; TDR, UNICEF/UNDP/World Bank/WHO Special Programme for Research and Training in Tropical Diseases; Kamuzu University of Health Sciences, Lilongwe, Malawi; School of Nursing and Rehabilitation, Shandong University, China; Department of Family of Medicine and Primary Care, School of Clinical Medicine, Faculty of Health Sciences, University of the Witwatersrand, Johannesburg, South Africa; Social Innovation in Health Initiative (SIHI) Indonesia at the Center for Tropical Medicine, Faculty of Medicine, Public Health, and Nursing, Universitas Gadjah Mada, Yogyakarta, Indonesia; Precise Consultancy and Training Service (Abu Dhabi UAE); Uppsala University, SIHI hub, Sweden; Division of Management and Community Pharmacy, Department of Pharmaceutics, Faculty of Pharmacy, Universitas Gadjah Mada, Yogyakarta, Indonesia; Because Stories, Storytelling and Communication Agency, South Africa; Hospital Escuela, Tegucigalpa, Honduras; Instituto de Enfermedades Infecciosas y Parasitología Antonio Vidal, Tegucigalpa, Honduras; Social Innovation in Health Initiative - Philippines Hub, Philippines; Department of Epidemiology and Environmental Health, University at Buffalo, State University of New York, NY, USA; Makerere University School of Public Health - Department of Community Health and the SIHI Uganda; Institute of Health Policy and Development Studies, National Institutes of Health of the University of the Philippines Manila, Philippines; Global Health & Innovation Lab, Faculty of Health Sciences, Western University, London, Ontario, Canada; African Centre for Innovation & Leadership Development, Abuja, Nigeria; Centre for Health and Sustainability, Department of Women’s and Children’s Health, Uppsala University, Sweden; Department of International Clinics, Shanghai Children’s Medical Center, Shanghai Jiao Tong University School of Medicine, China; University of North Carolina at Chapel Hill - Project China, USA; Department of Health Behavior,Gillings School of Global Public Health, University of North Carolina at Chapel Hill, USA

## Abstract

**Background:** Social innovation in health refers to the community-engaged process that connects health improvement and social change. The aim of this study was to develop a consensus statement on core learning competencies in social innovation in health and pilot them as part of a participatory training workshop.

**Methods and Findings:** A modified Delphi Process aggregating data from a scoping review, global open call, and participatory process was organized. Participants were recruited from low, middle, and high-income countries with a range of social innovation experiences. Statements focused on social innovation in health core competencies for learning. Consensus was determined using the RAND/UCLA Appropriateness method.

After expressing interest in the project, 68 individuals received the survey link. 46 participants completed the first survey, and 34 completed the second survey. All 28 statements reached consensus, and based on the results of this first survey, some statements were added, amended, and merged to reach 30 consensus statements in the second survey. Competencies were categorized into skills, mindsets, and knowledge. Some competencies reached higher levels of agreement than others. This included community engagement, which can leverage the collective knowledge and problem-solving abilities of a diverse group of individuals to tackle complex challenges; social entrepreneurship skills such as business model knowledge, securing funding, team building, and knowledge of intersectional issues and health inequities.

Several learning competencies were then piloted as eight one-hour online workshops, which assessed the feasibility of developing them through online open-access social innovation training sessions. After completing the workshops, 137 participants completed a survey, and most participants reported a significant improvement across six competencies.

**Conclusion:** The results from this study will inform the development of a WHO/TDR conceptual framework for teachers and learners in social innovation in health.

**Author Summary:** **Why was this study done?**

This study was undertaken to develop a consensus statement on core learning competencies in social innovation in health and pilot them as part of a participatory training workshop.

**What did the researchers do and find?**

Some of the core competencies that reached high levels of agreement amongst the international panel included community engagement, which can leverage the collective knowledge and problem-solving abilities of a diverse group of individuals to tackle complex challenges; social entrepreneurship skills such as business model knowledge, securing funding, team building, and knowledge of intersectional issues and health inequities.

**What do these findings mean?**

These findings are important for fostering social innovation in health training programmes and will inform the development of a WHO/TDR conceptual framework.

## Background

Social innovation in health is a community-engaged process that connects health improvement and social change [1]. Social innovation can create more effective, sustainable, and efficient solutions to serve the health needs of communities. Centring local communities to identify and develop innovative solutions has a wide range of benefits, and teaching core social innovation competencies can empower communities and other stakeholders to innovate for better health in their localities [2].

There is uncertainty about core competencies of social innovation in health that could be considered in training programs. While numerous social innovation education programs and training resources exist, content and methods vary significantly. The majority of learning resources have been developed for high-income settings [3]. The Special Programme for Research and Training in Tropical Diseases (TDR) and the Social Innovation in Health Initiative (SIHI) have highlighted the need for more social innovation training to catalyse and support the development of novel health solutions and build a pipeline of social innovation researchers [4].

The WHO uses consensus methods to develop normative guides for policymakers, national public health agencies, and others [5]. In 2023, TDR (the UNICEF/UNDP/World Bank/WHO Special Programme for Research and Training in Tropical Diseases) and SIHI indicated a need for consensus on social innovation in health learning competencies to be considered in training and further resources. This study addresses the critical gap in social innovation in health learning by establishing consensus on core competencies and piloting them through participatory workshops.

## Methods

### Modified Delphi Process

To develop consensus, this study used a modified Delphi, an iterative process of consolidating expert opinions into group consensus [6]. It was initially used in the 1950s to predict the impact of technology on society [7]. Modified Delphi approaches have been used across many health fields, especially in areas where there is a lack of evidence on best practices [8]. The RAND/UCLA Appropriateness method is a modified Delphi that involves iterative surveying and then group feedback sessions after the completion of the first survey [9]. This study used a two-round modified Delphi, supplemented by teleconference discussions **(Figure 1).**

**Figure 1:**
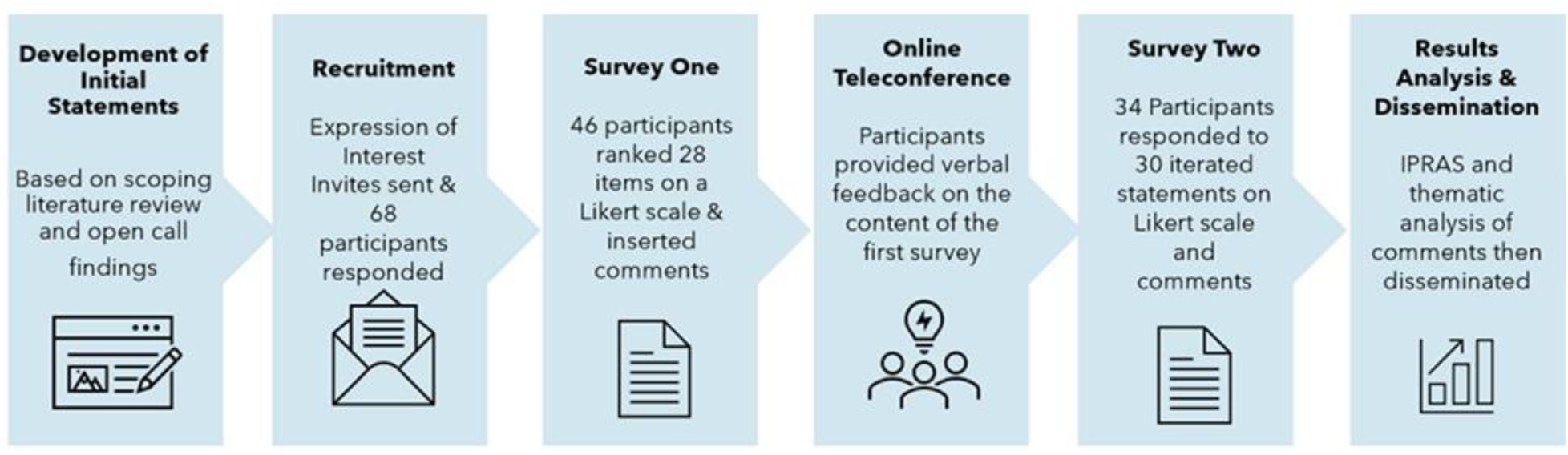
Steps in Consensus Building Process

Ethical approval was received in 2024 from the London School of Hygiene and Tropical Medicine, United Kingdom. There were no potential risks or harms to participants identified as questionnaires did not request sensitive information and participants were not deemed to be a vulnerable population. All participation was voluntary with informed consent. Raw data will be deleted one year after the completion of the study.

### Survey Development

The initial development of statements was informed by the results of a scoping literature review and a crowdsourcing open call conducted by Social Entrepreneurship to Spur Health (SESH) and SIHI. The open call was a structured crowdsourcing way of soliciting community feedback. The themes identified during the open call and scoping review included a mindset, which was further described as items based on empathy building, resilience, and adaptability. The skills grouping included communication, advocacy, partnership building, participatory research, and user-centered design, while the knowledge-based competencies contained intersectionality, advocacy, and ethical-related themes (**Figure 1).**

### Recruitment

A total of 113 members from the SIHI network, corresponding authors from the social innovation in health learning competency scoping review, and academics with a research interest in social innovation in health were invited to the study. Participants were emailed a link to a short survey hosted on the JotForm platform that requested contact details and basic demographic information. Inclusion criteria for participants included the ability to complete questionnaires in English, internet access, and a previous track record in social innovation in health activities. Participation was voluntary, and no incentives were offered.

### Round One

The first survey took approximately ten minutes to complete. Participants had two weeks to complete the survey in early June 2024, and a reminder email was sent five days prior to the deadline. The consent was embedded in the form. Demographic information such as country of birth, education level, gender, and specific details on their experience in social innovation were collected. The questionnaire contained 28 statements on core competencies. Each statement was presented initially with a definition, followed by a statement referring to social innovation in health. The panellists were asked to rank their level of agreement on a scale of 1-9, with 9 corresponding to strongly agree, 7 to agree, 5 to neutral, 3 to disagree, and 1 to strongly disagree. Each statement was followed by a Likert scale and a comment box where participants could provide feedback and propose statement edits.

### Teleconference Session

Participants were then invited to provide verbal feedback on the first survey and discuss statements on Zoom teleconference calls. The session was repeated two different times to facilitate attendance in different time zones. There was discussion on statements that had conflicting comments from the first survey, and then time was allocated for participants to suggest new competencies and propose iterations.

### Round Two

The second survey was developed, based on the analysis of the results of the initial survey and teleconference. The survey contained revised statements and definitions, along with some additional statements. Similarly to round one, each statement was followed by both a Likert scale and a comment box. Participants had the opportunity to suggest useful open-access resources and provide insights on how learning content delivery could be tailored to meet the needs of resource-limited contexts.

### Analysis

The median, interquartile range, participants’ ratings compared to group ratings, and frequencies for each item were calculated for all statements in both surveys.

The Inter-percentile Range Adjusted for Symmetry (IPRAS), derived from the RAND/UCLA appropriateness method, was used to assess the level of agreement among participants based on their Likert scale responses. IPRAS measures asymmetry in responses across the 9-point Likert scale using a calculation incorporating the inter-percentile range (30^th^ and 70^th^ centile), the median score, and a correction factor for asymmetry. This approach, applied to each rated statement, was developed as an alternative to relying on arbitrary cut-offs [10] [11]. The agreement was defined as per RAND definitions and assessed where the median falls [12]. A disagreement index (DI) below 1 indicated consensus, with a median score of 7-9 signifying agreement and 1-3 indicating disagreement. A thematic analysis of the comments was also conducted to identify aspects of statements for iteration and new statement additions to the survey. Themes that were repeatedly highlighted by different participants were included in the next iteration.

After data collection and analysis, all participants received a note of gratitude for their participation as well as feedback with summarised findings from the questionnaire and consensus statement. ACCORD (Accurate Consensus Reporting Document) was used to report study findings [13].

### Pilot Online Workshop

Twelve core competencies were then piloted as a training workshop series. From Jun 4 through Jul 23, 2024, an online training workshop on social innovation in health (60 minutes per week) was organized by SESH (Social Entrepreneurship to Spur Health) and SIHI. The workshop series consisted of eight online sessions. [14] Workshops covered the following topics: Community engagement, user-centered design and crowdsourcing (week 1), social innovation theories and frameworks and community engagement (week 2), human-centered design for health and empathy building (week 3), intersectional themes such as social determinants of health and health disparities (week 4), co-creation and community engagement (week 5), human-centered design (week 6), leadership and building sustainable innovations (week 7), pitching and storytelling for social innovations (week 8).

This component of the study was deemed not to be human subject research which was approved by the University of North Carolina Institutional Review Board. Data was de-identified and will be deleted one year after completion of the study.

Baseline assessments gathered information on participants’ sociodemographic characteristics, while self-administered exit surveys on the final day evaluated participants’ experiences and the effectiveness of the training. Participants were also asked to assess the overall workshop series and each session individually, focusing on their social innovation knowledge and skills, as well as their confidence in applying these concepts after the workshop. The Mann-Whitney U test was used to estimate changes by comparing self-assessed pre-workshop measures (i.e., reflecting on their knowledge and skills before starting the workshop) with post-workshop measures. All data were analyzed using GraphPad Prism 8.0 statistical software (GraphPad Software Inc., La Jolla, CA, USA).

## Results

A total of 68 individuals expressed interest in participating in the Delphi process in June 2024. Forty-five (66.6%) of those identified as women. Twenty-four countries from all WHO regions were represented. Nigeria, the Philippines, Sweden, and the United States of America had the highest representation from individual countries, with at least five people expressing interest in participating. The response rate for the first survey was 46/68 (67.6%). Participants resided in 18 different countries. Thirty-three (71.7%) were female, and there were various experiences in social innovation and education level, as depicted in **Table 1**.

**Table 1:**
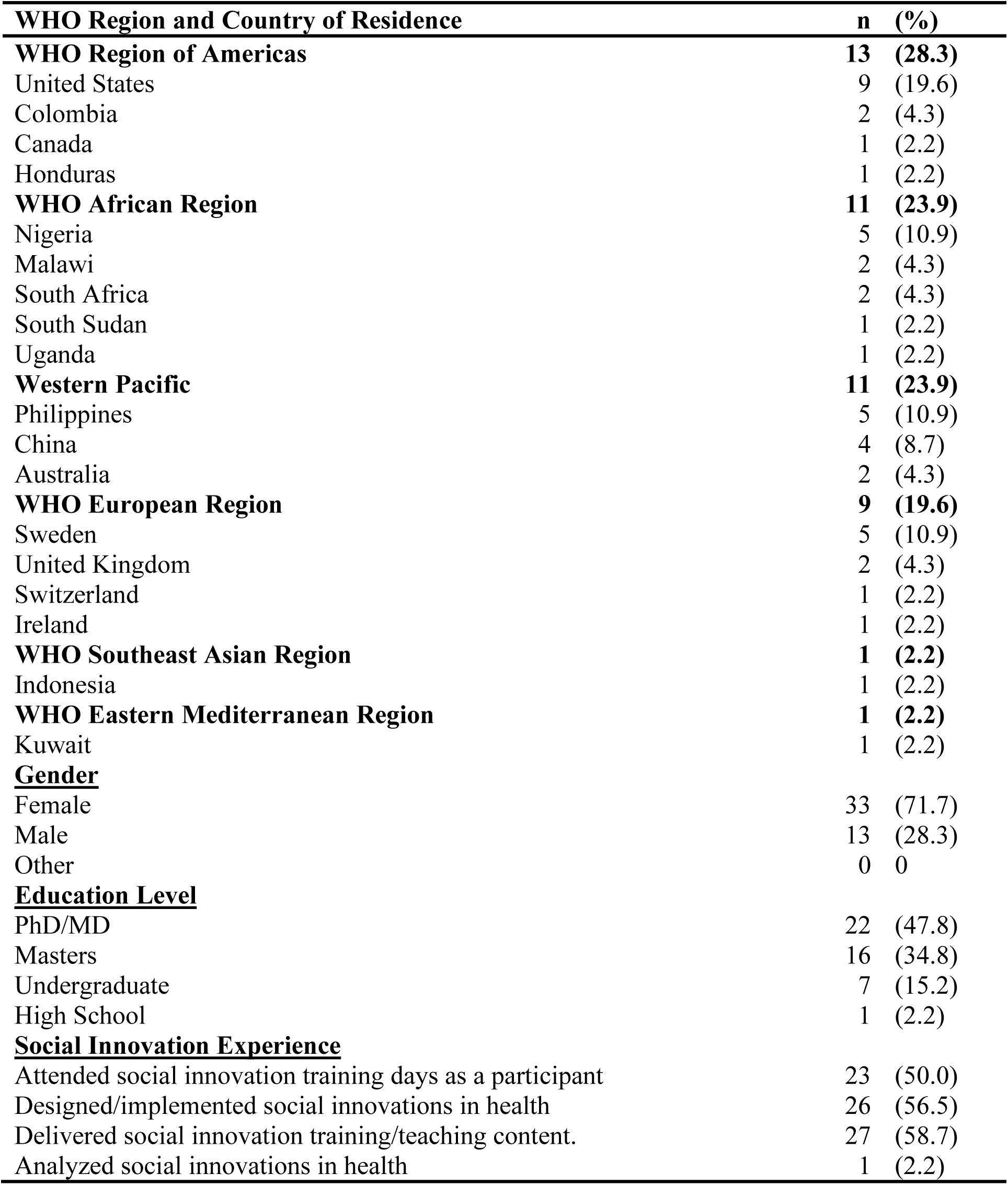
Demographic Details of Participants who completed the first Survey on Learning Competencies in Social Innovation in Health.

| **WHO Region and Country of Residence** | **n** | **(%)** |
| --- | --- | --- |
| **WHO Region of Americas** | **13** | **(28.3)** |
| United States | 9 | (19.6) |
| Colombia | 2 | (4.3) |
| Canada | 1 | (2.2) |
| Honduras | 1 | (2.2) |
| **WHO African Region** | **11** | **(23.9)** |
| Nigeria | 5 | (10.9) |
| Malawi | 2 | (4.3) |
| South Africa | 2 | (4.3) |
| South Sudan | 1 | (2.2) |
| Uganda | 1 | (2.2) |
| **Western Pacific** | **11** | **(23.9)** |
| Philippines | 5 | (10.9) |
| China | 4 | (8.7) |
| Australia | 2 | (4.3) |
| **WHO European Region** | **9** | **(19.6)** |
| Sweden | 5 | (10.9) |
| United Kingdom | 2 | (4.3) |
| Switzerland | 1 | (2.2) |
| Ireland | 1 | (2.2) |
| **WHO Southeast Asian Region** | **1** | **(2.2)** |
| Indonesia | 1 | (2.2) |
| **WHO Eastern Mediterranean Region** | **1** | **(2.2)** |
| Kuwait | 1 | (2.2) |
| **<u>Gender</u>** |  |  |
| Female | 33 | (71.7) |
| Male | 13 | (28.3) |
| Other | 0 | 0 |
| **<u>Education Level</u>** |  |  |
| PhD/MD | 22 | (47.8) |
| Masters | 16 | (34.8) |
| Undergraduate | 7 | (15.2) |
| High School | 1 | (2.2) |
| **<u>Social Innovation Experience</u>** |  |  |
| Attended social innovation training days as a participant | 23 | (50.0) |
| Designed/implemented social innovations in health | 26 | (56.5) |
| Delivered social innovation training/teaching content. | 27 | (58.7) |
| Analyzed social innovations in health | 1 | (2.2) |

All of the 28 statements, all met the threshold for consensus to varying degrees. A total of 25 of 28 (89.2%) statements had a median Likert rating score of >8 and a Disagreement Index <0.3, indicating strong agreement. Statements that had very strong agreement included communication-based competencies and competencies based on intersectional topics. Other items, such as manuscript writing for scientific publication, saw diverging perspectives among participants. 28 of 46 (60%) participants entered comments to at least one statement - demonstrating a high level of participant engagement. Based on this commentary, the definitions of 26 statements were elaborated for the second round of the survey. Four statements with common themes were merged, and three new items were added, including proposed competencies in the area of research skills, navigating regulatory pathways, and definitions and frameworks as a practical tool.

### Teleconference Sessions

A total of 34 participants joined teleconference sessions. Different methods of learning were discussed, such as classroom-based teaching styles, self-directed learning, and experiential learning. Participants gave feedback that research skills such as literature appraisal were important to include in the subsequent iteration, along with an emphasis on more participatory approaches to achieve social innovation as competencies.

### Second Survey

The response rate was 36/68 (52.9%) for the second survey. One participant was excluded because they did not have experience in social innovation. Twenty-four (68.6%) participants were women, and there was representation from 20 countries. The majority (77.1%) of participants had either a PhD, MD or master’s degree. Participants had varying experience in social innovation in health among participants, including teaching and learning about social innovation and delivering and implementing innovations. The threshold for consensus was reached in all 30 statements in the second survey. In general, comments varied in sentiment, with the majority expressing agreement with the statements.

### Competencies

We identified 30 competencies, categorized into three groups: 3 related to mindset/attitude, 17 focused on skills, and 10 based on knowledge-related competencies **(Figure 2).**

**Figure 2:**
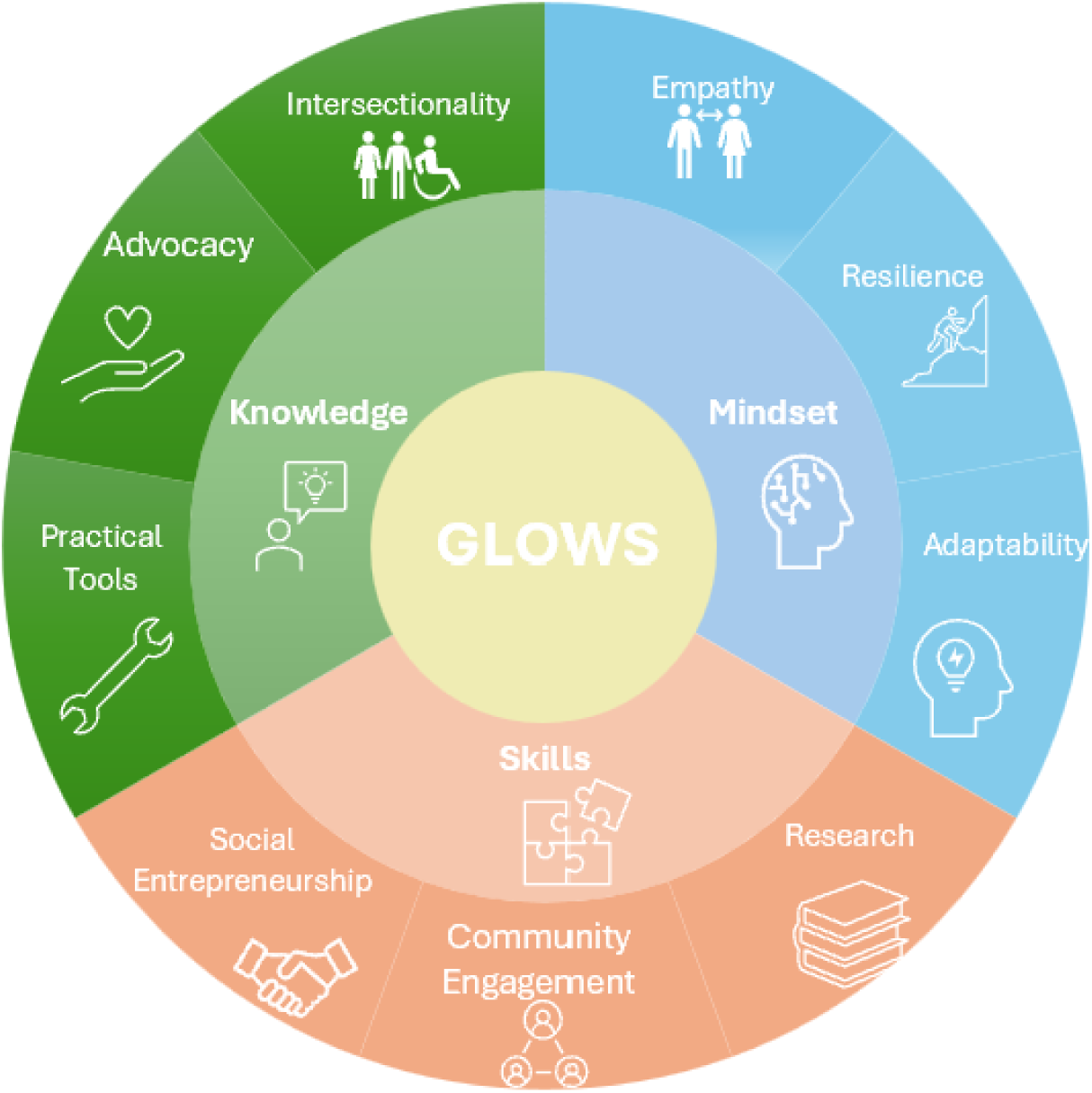
Core Competencies including Skills, Knowledge, and Mindsets Identified from the Consensus Building Process

Empathy, adaptability, and resilience were all mindset-based competencies that were included. Each had a median Likert ranking of at least 8, indicating strong agreement, and 34/35 (97.1%) of participants agreed to varying degrees with the statements. While, overall, participants felt that having the right mindset should be included as an aim for learners to enable success in social innovation, numerous participants felt that one’s mindset is an intrinsic trait that is difficult to teach. On the other hand, some felt that it is feasible to develop mindsets.

There was also strong agreement for most of the skill-based competencies. All communication-based competencies (such as community engagement, storytelling, and pitching) received high ratings. Storytelling and pitching were identified as necessary to gain support, mobilize resources, and inspire others to action. Community engagement leverages the collective knowledge, creativity, and problem-solving abilities of a diverse group of individuals to tackle complex challenges. Social entrepreneurship skills such as business model knowledge, securing funding, team building, campaigning, and developing partnerships were also deemed to be important for social innovation.

Research skills were another important competency identified. Some research skills such as community based participatory research reached very strong levels of agreement resulting in a DI of 0.13, while others slightly weaker agreement such as manuscript writing manuscript writing for scientific publication had a disagreement index (DI) of 0.65.

Generally, participants indicated that social innovators should accomplish a broad range of skills, but it was acknowledged in many responses that some of these skill sets are not necessary in all cases. Having an awareness of these competencies and their importance is crucial for innovators to be able to identify the need for expertise in the area, which could then be outsourced.

Developing a knowledge base on intersectional issues, advocacy, ethics, and the practical tools that can be used in social innovation were all items that also met consensus. Intersectionality refers to the complex, cumulative way in which the effects of multiple forms of discrimination (such as racism, sexism, and classism) combine, overlap, or intersect, especially in the experiences of marginalized individuals or groups [15]. Knowledge of intersectionality in this context covered topics such as understanding health disparities, considering how social determinants of health influence these disparities, and navigating diverse cultural contexts respectfully. An understanding of these topics is key for alignment with social innovation values, which are community-centricity, co-creation, and a deep respect for local context. The median Likert rating was 9, with a very low disagreement index of 0.13 on all five statements relating to intersectional themes. This demonstrates participants’ agreement on the importance of learning about these issues to advance health equity through social innovation.

Knowledge of advocacy and ethics were also valuable competencies identified by the participant cohort. Commentary from several participants indicated that the inclusion of ethical frameworks and guidelines and the establishment of clear accountability measures were significant. Through the consensus-building process, advocacy tools were defined as including policy analysis and lobbying. Respondents expressed that such techniques are useful to influence decision-makers and shift their mindsets.

The final list of competencies is summarized in Table 2, with key themes highlighting community engagement, social entrepreneurship, and knowledge of intersectionality and health disparities.

**Table 2:**
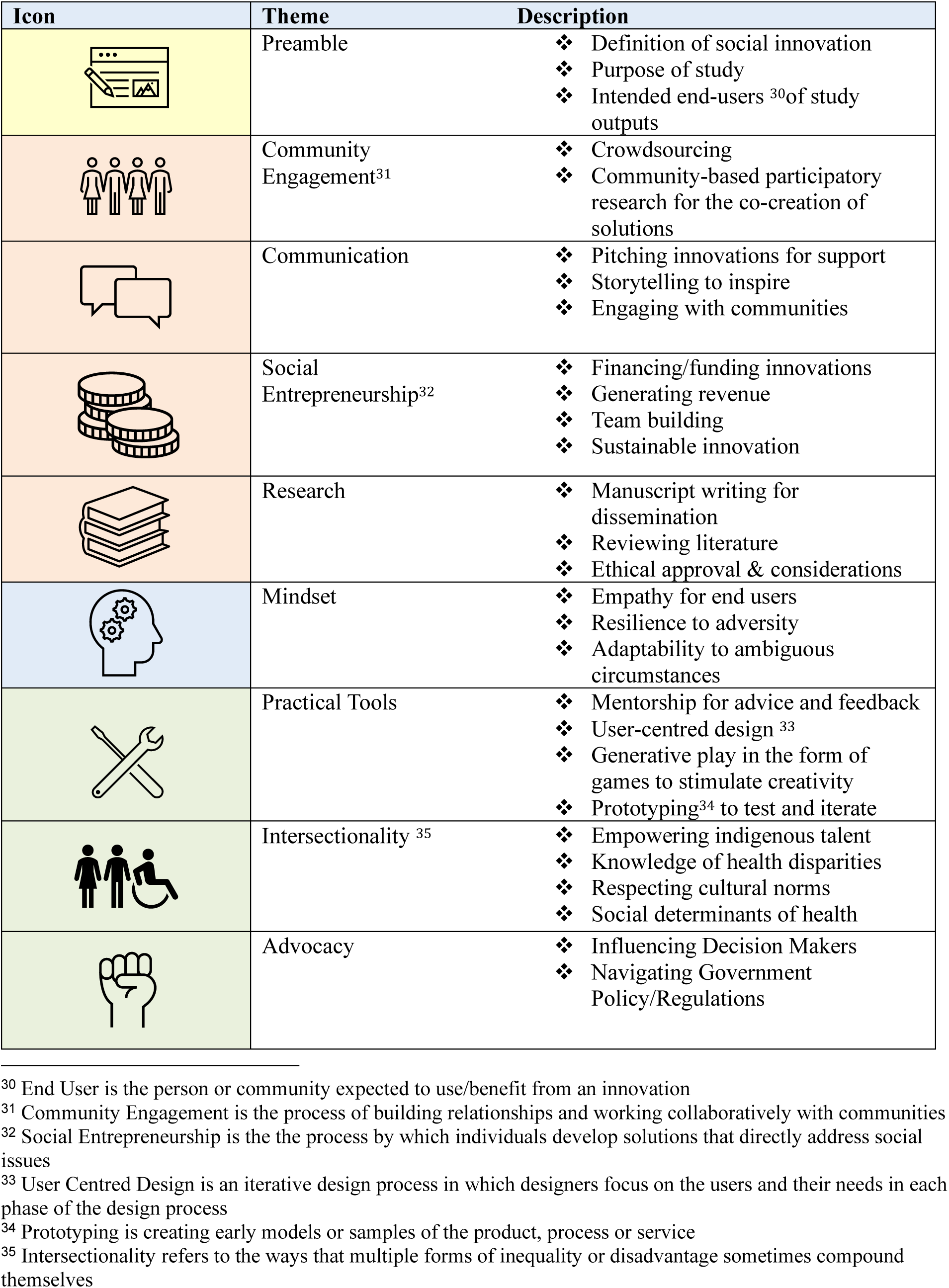
Summarized Learning Competency Statements, Second Survey.

### Learning Resources & Project Outputs

Contributors suggested open-access learning resources, which included a variety of tools such as videos, practical guides, free online courses, websites, and case studies. WHO/UNAIDS toolkit for community engagement, SESH grant-o-thon, Massive Open Online Courses, success stories, budget and proposal templates, and monitoring and evaluation framework were also highlighted. Repeated themes mentioned were the availability of open-access resources and the availability of material in local languages to promote health equity.

### Online Training workshop

A total of 137 participants completed the exit survey. Based on the final evaluation survey, most participants were from the Western Pacific Region (50.4%, n=69) and upper-middle-income countries (59.9%, n=82). About 70.1% of responders (n=96) were female, and a small number identified as belonging to a racial or ethnic minority group (10.2%, n=14). Over half (59.9%, n=82) were students, and nearly half (46.0%, n=63) had already held a master’s degree. Public health was the most common field of previous training (42.3%, n=58), with research (45.5%, n=25) being the primary job duty. In addition, our participants had an average of 9 ± 6.9 years of work experience. As for the post-workshop experiences, 52.6% (n=72) of the participants attended all eight workshop sessions. The workshop goals were mostly or completely met for 78.1% (n=107) of participants, and the majority rated the overall workshop series as excellent (67.9%, n=93). Nearly all participants (96.4%, n=132) expressed willingness to attend future workshops.

The training impact among participants was evaluated through a pre-and post-assessment of their self-reported social innovation knowledge and skills, using a scale from 1 (not at all knowledgeable) to 5 (very knowledgeable), as well as their confidence in applying these concepts post-workshop, ranging from “not confident at all” to “very confident”. Additionally, participants rated their competency in seven key learning areas on a scale from 1 (not competent at all) to 5 (very competent). Overall, participants increased their social innovation knowledge scores by 1.0 and improved their confidence by 1.0 points (**Figure 3**). Most participants reported a significant improvement, with a 1-point increase in learning competency across six areas after completing the workshop.

**Figure 3.**
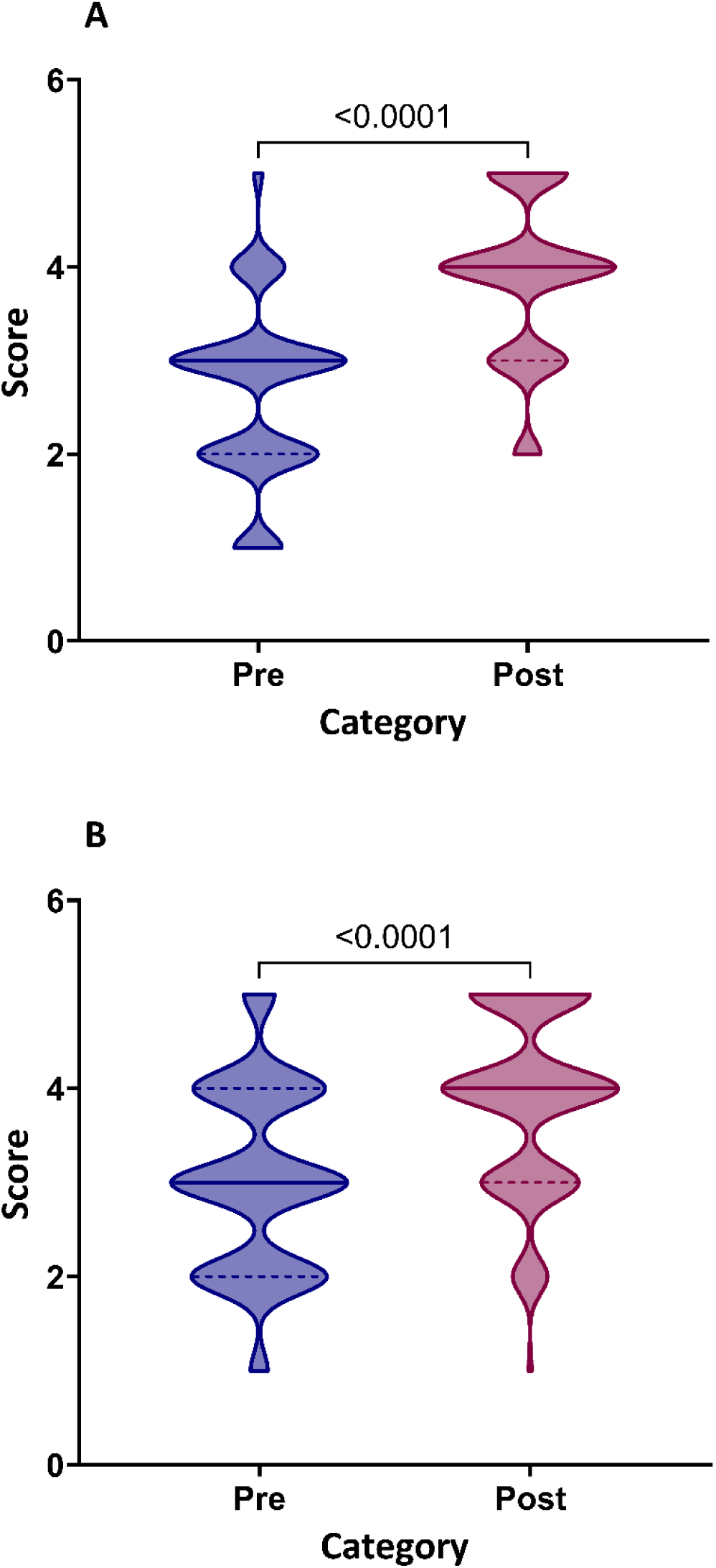
Overall enhancement of A. social innovation knowledge/skills and B. confidence in applying these concepts post-workshop. The violin width in Figure 3 represents the number of participants at a certain value. Solid lines indicate the median value, and dotted lines indicate the IQR. Statistical significance between groups was assessed using the Mann–Whitney U test.

In the context of what participants found the most useful in the workshop series, our analysis generated several themes: ‘developing social innovation core competencies’, ‘applying knowledge for real-world impact’, ‘embracing diverse perspectives and building collaborations’, ‘engaging in interactive and practical learning’, ‘offering mentorship and providing guidance’ ‘inspiring and motivating through shared experiences’. Specifically, in the theme of ‘developing social innovation core competencies’, participants highlighted how they had been exposed to the concept, knowledge, and skills of social innovations in health: “*I learned a lot about social innovations in health and how they can apply to tackle health and developmental challenges in a largely populated third world country like Nigeria*“; “*Create impactful, equitable, and sustainable health solutions by actively involving communities and leveraging their strengths*”. In addition, in terms of applying knowledge for real-world impact, some of our participants emphasized how they had benefitted and enhanced their capacity to address real-world challenges: “*The workshop’s practical tools…. directly enhanced my projects and boosted my confidence…*”; “*Applying theories and best practices … was invaluable*”.

## Discussion

This study identified and piloted learning competencies focused on social innovation in health. The findings from the consensus process and pilot training workshops will be valuable to those learning about social innovation who aim to deliver solutions to improve health, as well as those teaching social innovation courses. This expands the literature by focusing on learning about social innovation, capturing ideas from a diverse LMIC end-user group, and leveraging participatory methods critical for learning about social innovation.

Some of the competencies with the greatest consensus included community engagement, social entrepreneurship, and knowledge of intersectional issues. Community engagement was identified to be a crucial skill. It is well recognized as one of the main pillars of social innovation in the literature and is often contained within the definition of social innovation [16]. There are several mechanisms to engage communities, including community-based participatory research and crowdsourcing, which are recognized in this consensus statement [17].

Knowledge of intersectionality was also identified to be a core competency. There were five statements relating to intersectionality, all of which achieved high levels of agreement. Knowledge of intersectional issues is key to addressing health problems alongside related social determinants of health, and it has been consistently identified in the literature and open calls. Achieving an understanding of these intersectional topics is sometimes omitted from social innovation education; however, there are many teaching resources focused on intersectionality, and there is a growing body of literature linking intersectionality and social innovation [18] [19] [20].

Social entrepreneurship skills also were deemed to be important for learning about social innovation. Leveraging a network and collaborating with those with varied expertise and skills to build the right team are also essential skills in social entrepreneurship, and the majority of participants agreed that they were crucial [21]. There is some limited but growing literature emphasizing the importance of social entrepreneurship skills among health practitioners and communities, but additional resources are needed to help learners [22].

This study also has limitations. First, the modified Delphi methodology has been criticized for lacking reliability and reproducibility. We anticipated this and increased the rigor by using participatory approaches, recruiting diverse participants, and reporting according to the ACCORD consensus reporting guidelines. Second, our participants were more familiar with social innovation compared to other people. However, this consensus statement would be most relevant for teachers, professors, and others responsible for organizing social innovation courses. Third, we did not examine all core competencies in the pilot. However, we did examine 12 of the critical learning competencies identified in the Delphi study.

Our data have implications for policy and practice. The data from this consensus statement can inform practical guides on social innovation learning. This can be widely distributed to ensure consistency in achieving social innovation learning competencies. Further evidence is needed on the best ways to support learners in achieving the outlined competencies. More open-access resources should be developed and made available targeting these competencies. Furthermore, as most of the existing resources for learning about social innovation in health are in English, translation of key tools into local languages so that they are accessible to a more diverse range of communities.

To conclude, a two-round modified Delphi was conducted, examining the core learning competencies that should be achieved by those learning about social innovation in health. Participants were recruited to represent global social innovation activity, and statements were developed based on a scoping review and international open-call findings. Consensus was achieved on all items through a participatory approach. Mindset, skills, and knowledge in the appropriate domains contribute to success in social innovation. Innovative ways of learning should be considered to achieve important learning competencies, which may include structured content, self-directed learning, and experiential learning. The findings from this study will inform the development of a practical framework. This will be important in capability building in social innovation education to solve complex global health challenges.

## Data Availability

All data produced in the present study are available upon reasonable request to the authors

## Acknowledgements

We would like to thank all participants in the open call. The work received support from TDR, the Special Programme for Research and Training in Tropical Diseases co-sponsored by UNICEF, UNDP, the World Bank and WHO. TDR is able to conduct its work thanks to the commitment and support from a variety of funders. These include our long-term core contributors from national governments and international institutions, as well as designated funding for specific projects within our current priorities. For the full list of TDR donors, please visit TDR’s website at: https://www.who.int/tdr/about/funding/en/ TDR receives additional funding from Sida, the Swedish International Development Cooperation Agency, to support SIHI.

## Supporting Materials

**S1 Table.** Description of each session of the Social Innovation in Health Mid-year Training Workshop.

| Title |  | Summary | Learning Competencies |
| --- | --- | --- | --- |
| Week 1 | Introduction to Participatory Health Research Methods, Emphasizing Designathons | Participatory research approaches like community advisory boards, co-creation, and design-a-thons provide an avenue for formative assessments and serve as the basis for collaboration with end-users in identifying solutions to gaps in implementation and improving efficiency. | • Community Engagement<br>• User-centred design<br>• Crowd-sourcing |
| Week 2 | Social Innovation in Health: Introduction to Social Innovation Theory and Knowledge, Attitudes, Practices, and Behaviours (KAP-B) Framework | This session introduces the audience to the concepts and applications of social innovation theories in health. The learning objectives of this session include: A. To define social innovation in health and its implications for inclusivity and community engagement. B. To explain social innovation theories, such as diffusion of Innovations Theory and Social Capital Theory, and their relevance in creating social value and systemic change. C. To explain the KAP-B Framework and its application in addressing social issues. | • Community Engagement<br>• Theories and Frameworks |
| Week 3 | Introduction to Human-Centered Design for Health | This session focuses on the principles of human-centered design and its application through an online interactive workshop. The session will briefly cover basic concepts, such as the phases of human-centered design, before guiding participants in a series of activities to apply what they have learned to a case vignette provided. This session aims to equip participants with knowledge, tools, and resources related to human-centered design that they can use for their work in health and social innovation. | • Human Centred Design<br>• Empathy |
| Week 4 | Applying an Intersectional Gender Lens | We are all shaped by different aspects of identity, such as race, gender, and social class. | • Intersectionality<br>• Social Determinants of Health |
|  | on Social Innovations in Health: An Introductory Module | Intersectionality helps us understand how these layers interact and influence our experiences. This session will focus on gender as a key entry point into intersectional analysis, examining how gender power relations intersect with other social factors to impact lives and create diverse needs and experiences. Through this one-hour interactive session, participants will gain an overview of the importance of gender frameworks in research and learn how to apply an intersectional gender framework in social innovations in health. Understanding these intersections is essential for future gender-sensitive and transformative research. | • Navigating Diverse Cultural Contexts<br>• Health Disparities |
| Week 5 | Co-Creation: An Innovative Communal Approach in Addressing Social Health Challenges | This workshop session is designed for individuals and groups interested in using a Living Lab Bayanihan- a communal approach focused on co-creation and multi-stakeholder collaboration in addressing social health challenges. This session will include interactive activities and practical strategies for Problem Identification and the Importance of User/People and Stakeholder Engagement. | • Community Engagement |
| Week 6 | Democratizing Applied Behavioral and Decision Sciences (BeSci) to Combat MMR Vaccine Hesitancy through Behavioral Design Sprints | AHA! Behavioral Design® (AHA! BD) pioneers the local application of Behavioral Science (BeSci), offering a Global South interpretation. Integrating Behavioral and Cultural Insights (BCI) into health promotion expands BeSci beyond nudges to tackle complex public health and sustainability challenges. This led to the Behavioral Design Sprint (BeDesign Sprint™), turning otherwise technical BCI into practical and actionable health solutions co-created with the community themselves. The session showcases a case study of this approach and an interactive condensed simulation of AHA! BD's | • Human-centred Design |
|  |  | BeDesign Sprint™ for audiences to participate in. |  |
| Week 7 | Embedding Social Innovation to Strengthen Health Systems and Leadership | This session explores how social innovation strengthens health systems by addressing current issues and developing key leadership competencies. It includes methodologies, tools, and success stories, such as the comprehensive intervention in La Guajira, Colombia, and the case of the Zika children. Leadership strategies that promote inclusion and collaboration among multiple stakeholders are highlighted. Additionally, participatory activities are conducted to prioritize health problems and analyze the perspectives from which leadership is exercised. The importance of social innovation in improving the efficiency and effectiveness of health interventions is emphasized, ensuring that the prioritized needs of the population are met. | • Leadership/team management<br>• Sustainable innovations |
| Week 8 | From Idea to Impact: Mastering the Pitch for Social Innovations | You might have a groundbreaking social innovation, but it may not reach its full potential unless you convince others to believe in it through a compelling pitch. Participants will be introduced to key elements and strategies for an excellent pitch to effectively communicate a social innovation with a broad range of communities, especially people with lived experience and potential partners. In addition, through case studies, you will be exposed to concepts that can enhance your entrepreneurial skills to raise funds for social innovation and innovative financing. | • Pitching<br>• Storytelling<br>• Fund Raising |

**S2 Table:** Summarized Consensus Items and IPRAS ratings.

| # | Statement | Disagreement Index | Median | Consensus A= Agreement |
| --- | --- | --- | --- | --- |
| 1 | **Preamble:** social innovation definition | 0.29 | 8 | A |
| **Mindset** |  |  |  |  |
| 2 | Developing **empathy** so that the beneficiaries' needs are understood | 0.13 | 8 | A |
| 3 | Developing **resilience** should be an important focus area for learners | 0.13 | 8 | A |
| 4 | **Adaptability** to rapidly iterate and respond to local contexts | 0.13 | 9 | A |
| **Skills: Communication** |  |  |  |  |
| 5 | **Pitching skills** are an important competency | 0.13 | 8 | A |
| 6 | **Engaging communities** is a key skill to develop | 0.1 | 9 | A |
| 7 | **Storytelling** is an important skill | 0.13 | 8 | A |
| **Intersectionality** |  |  |  |  |
| 8 | Lessons on navigating diverse cultural contexts are essential | 0.13 | 9 | A |
| 9 | Understanding **health disparities** related to intersectional issues | 0.13 | 9 | A |
| 10 | Skills to identify **social determinants** that cause health disparities | 0.13 | 9 | A |
| 11 | Prioritizing indigenous talents and wisdom is an important skill | 0.13 | 9 | A |
| 12 | Engage with community members, conduct needs assessments, and gather insights into the target population is important | 0.13 | 9 | A |
| **Evaluation** |  |  |  |  |
| 13 | Teaching **evaluation** of social innovation/impact assessment | 0.13 | 8 | A |
| **Dissemination** |  |  |  |  |
| 14 | **Manuscript writing** is a key skill to learn | 0.65 | 7 | A |
| 15 | **Effective media use** (incl. social media) for information dissemination | 0.19 | 8 | A |
| **Entrepreneurship** |  |  |  |  |
| 16 | Navigating **funding pathways** such as grant writing, securing investment and business model knowledge is an important skill | 0.29 | 8 | A |
| 17 | Lessons on building and **managing a team** are important | 0.29 | 8 | A |
| 18 | Teaching about creating sustainable innovation is important | 0.13 | 9 | A |
| 19 | Lessons in **network management** are important | 0.29 | 8 | A |
| **Ethics and Advocacy** |  |  |  |  |
| 21 | **Ethical considerations** and pathways should be included | 0.16 | 9 | A |
| 22 | **Advocacy tools** should be taught to overcome barriers in health systems | 0.13 | 9 | A |
| **Practical Tools** |  |  |  |  |
| 20 | **Crowdsourcing** to identify social innovations is a useful tool to teach | 0.23 | 8 | A |
| 23 | **User-centred design** is important to deliver comprehensive training | 0.13 | 8 | A |
| 24 | **Mentoring** is an important tool to achieve core learning competencies. | 0.29 | 8 | A |
| 25 | **Prototyping** is important in allowing innovators to test & iterate their ideas | 0.29 | 9 | A |
| 26 | Using **generative play** can be a useful tool | 0.29 | 8 | A |
| 27 | Definitions, theories, frameworks, and case studies are all key tools | 0.26 | 8 | A |
| 28 | Navigating government policy, including regulatory landscapes | 0.29 | 8 | A |
| **Research** |  |  |  |  |
| 29 | **Research skills**, including conducting literature reviews | 0.37 | 7 | A |
| 30 | Community-based participatory research | 0.13 | 9 | A |

**S3: Expression of Interest Form** https://form.jotform.com/241321658160046

**S4: Survey One GLOWS: [Global Learning Opportunities Within Social Innovation](https://form.jotform.com/241242842128047) (jotform.com)**

**S5: Survey Two [Survey 2: Global Learning Opportunities Within Social Innovation](https://form.jotform.com/241745028024349) (jotform.com)**

**S6 Table:**
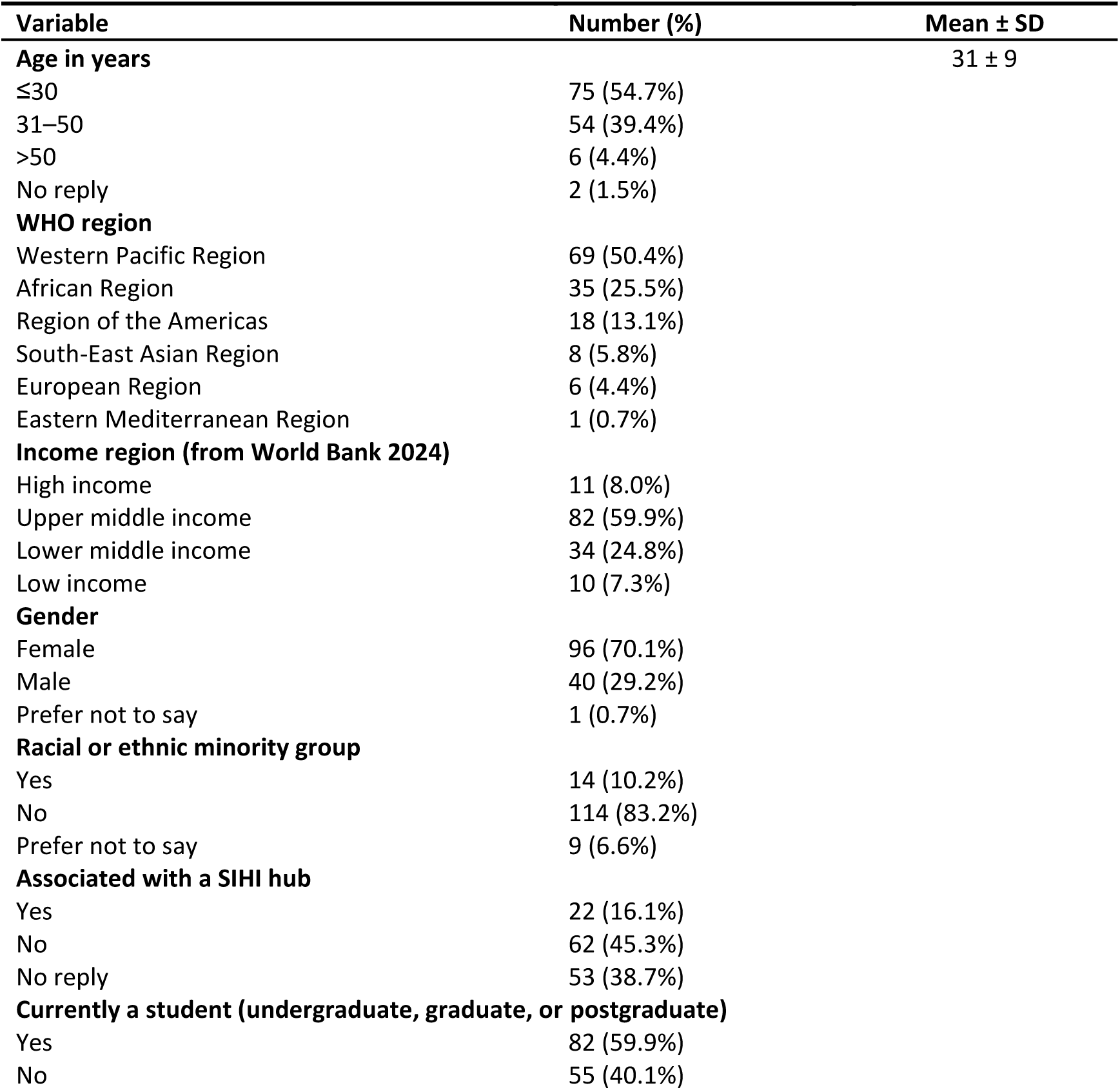
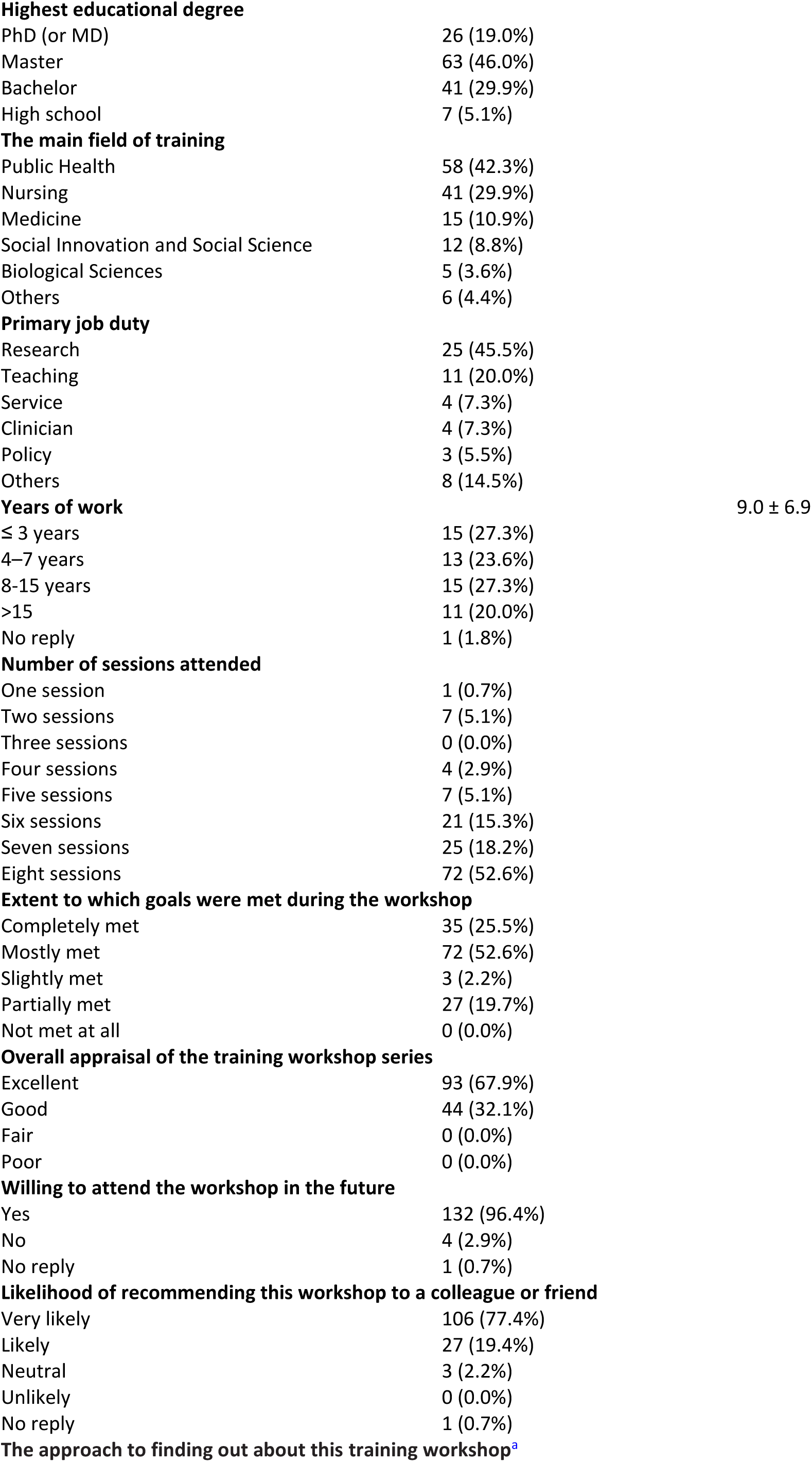
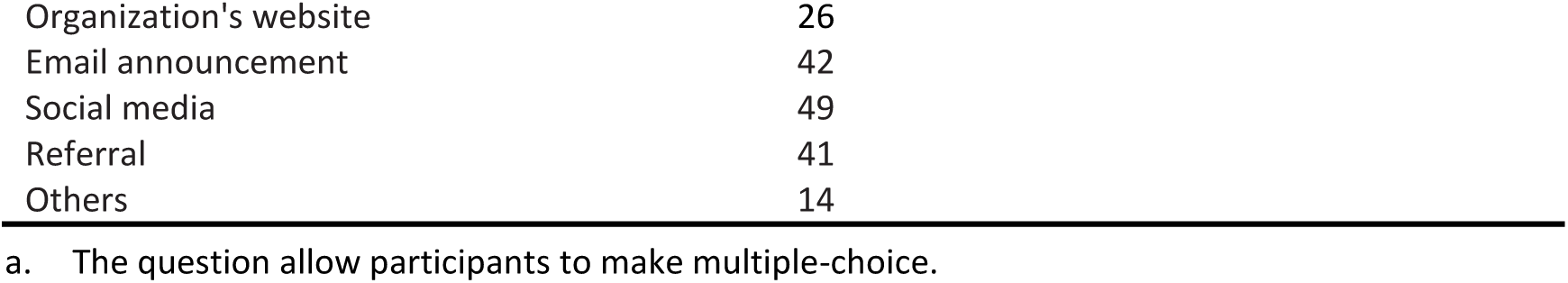
Demographic characteristics of participants attending the Social Innovation Mid-year Training Workshop held between June 2024 and July 2024 (N=137).

| Variable | Number (%) | Mean ± SD |
| --- | --- | --- |
| **Age in years** | | 31 ± 9 |
| $\leq 30$ | 75 (54.7%) | |
| 31–50 | 54 (39.4%) |  |
| >50 | 6 (4.4%) |  |
| No reply | 2 (1.5%) |  |
| **WHO region** |  |  |
| Western Pacific Region | 69 (50.4%) |  |
| African Region | 35 (25.5%) |  |
| Region of the Americas | 18 (13.1%) |  |
| South-East Asian Region | 8 (5.8%) |  |
| European Region | 6 (4.4%) |  |
| Eastern Mediterranean Region | 1 (0.7%) |  |
| **Income region (from World Bank 2024)** |  |  |
| High income | 11 (8.0%) |  |
| Upper middle income | 82 (59.9%) |  |
| Lower middle income | 34 (24.8%) |  |
| Low income | 10 (7.3%) |  |
| **Gender** |  |  |
| Female | 96 (70.1%) |  |
| Male | 40 (29.2%) |  |
| Prefer not to say | 1 (0.7%) |  |
| **Racial or ethnic minority group** |  |  |
| Yes | 14 (10.2%) |  |
| No | 114 (83.2%) |  |
| Prefer not to say | 9 (6.6%) |  |
| **Associated with a SIHI hub** |  |  |
| Yes | 22 (16.1%) |  |
| No | 62 (45.3%) |  |
| No reply | 53 (38.7%) |  |
| **Currently a student (undergraduate, graduate, or postgraduate)** |  |  |
| Yes | 82 (59.9%) |  |
| No | 55 (40.1%) |  |
| **Highest educational degree** |  |  |
| PhD (or MD) | 26 (19.0%) |  |
| Master | 63 (46.0%) |  |
| Bachelor | 41 (29.9%) |  |
| High school | 7 (5.1%) |  |
| **The main field of training** |  |  |
| Public Health | 58 (42.3%) |  |
| Nursing | 41 (29.9%) |  |
| Medicine | 15 (10.9%) |  |
| Social Innovation and Social Science | 12 (8.8%) |  |
| Biological Sciences | 5 (3.6%) |  |
| Others | 6 (4.4%) |  |
| **Primary job duty** |  |  |
| Research | 25 (45.5%) |  |
| Teaching | 11 (20.0%) |  |
| Service | 4 (7.3%) |  |
| Clinician | 4 (7.3%) |  |
| Policy | 3 (5.5%) |  |
| Others | 8 (14.5%) |  |
| **Years of work** |  | 9.0 ± 6.9 |
| ≤ 3 years | 15 (27.3%) |  |
| 4–7 years | 13 (23.6%) |  |
| 8–15 years | 15 (27.3%) |  |
| >15 | 11 (20.0%) |  |
| No reply | 1 (1.8%) |  |
| **Number of sessions attended** |  |  |
| One session | 1 (0.7%) |  |
| Two sessions | 7 (5.1%) |  |
| Three sessions | 0 (0.0%) |  |
| Four sessions | 4 (2.9%) |  |
| Five sessions | 7 (5.1%) |  |
| Six sessions | 21 (15.3%) |  |
| Seven sessions | 25 (18.2%) |  |
| Eight sessions | 72 (52.6%) |  |
| **Extent to which goals were met during the workshop** |  |  |
| Completely met | 35 (25.5%) |  |
| Mostly met | 72 (52.6%) |  |
| Slightly met | 3 (2.2%) |  |
| Partially met | 27 (19.7%) |  |
| Not met at all | 0 (0.0%) |  |
| **Overall appraisal of the training workshop series** |  |  |
| Excellent | 93 (67.9%) |  |
| Good | 44 (32.1%) |  |
| Fair | 0 (0.0%) |  |
| Poor | 0 (0.0%) |  |
| **Willing to attend the workshop in the future** |  |  |
| Yes | 132 (96.4%) |  |
| No | 4 (2.9%) |  |
| No reply | 1 (0.7%) |  |
| **Likelihood of recommending this workshop to a colleague or friend** |  |  |
| Very likely | 106 (77.4%) |  |
| Likely | 27 (19.4%) |  |
| Neutral | 3 (2.2%) |  |
| Unlikely | 0 (0.0%) |  |
| No reply | 1 (0.7%) |  |
| **The approach to finding out about this training workshop<sup>a</sup>** |  |  |

|  |  |
| --- | --- |
| Organization's website | 26 |
| Email announcement | 42 |
| Social media | 49 |
| Referral | 41 |
| Others | 14 |

a. The question allow participants to make multiple-choice.

**S7: Figure.**
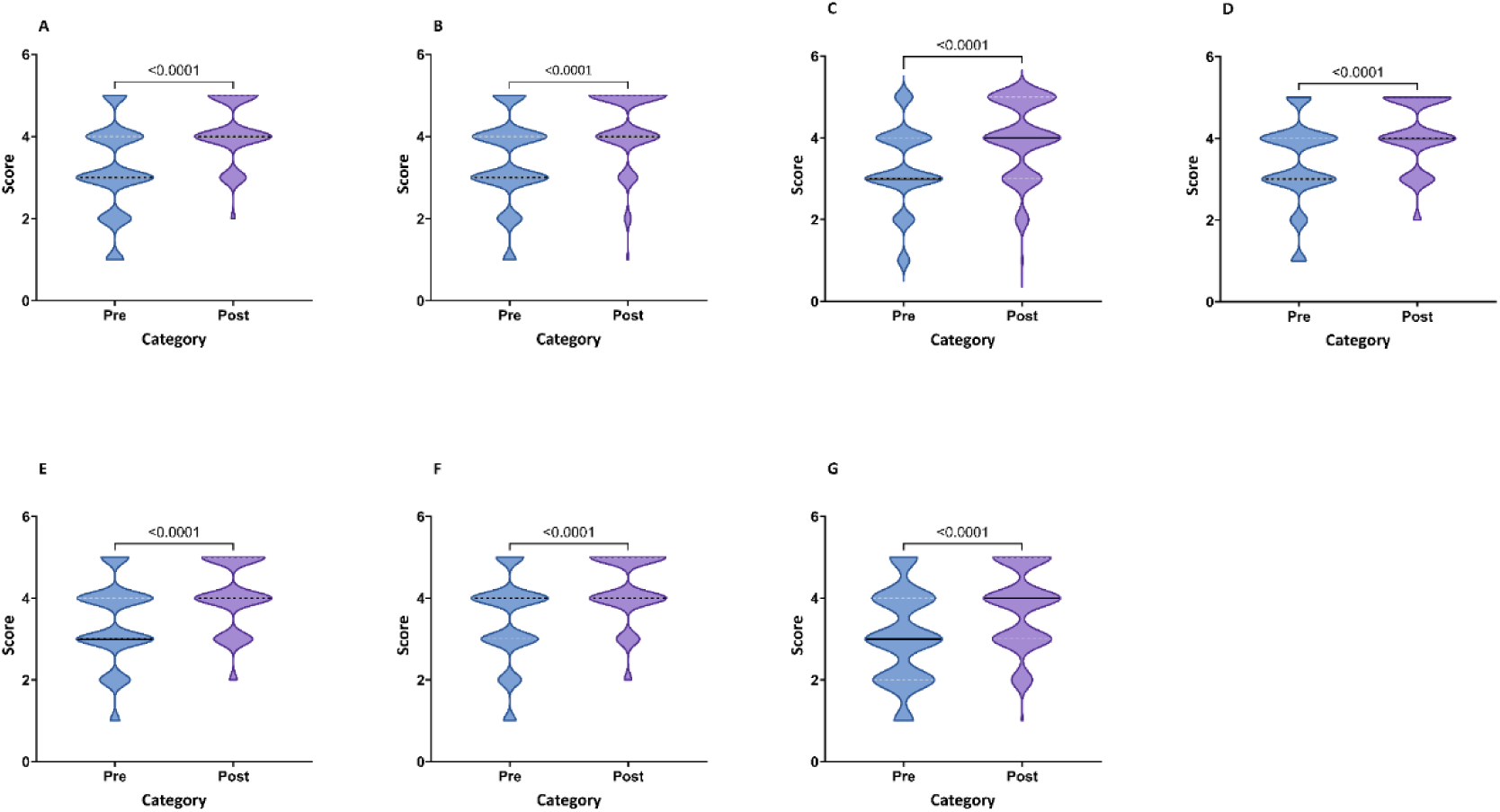
Enhanced social innovation in health knowledge/skills across seven competencies: A. Understanding health disparities and their roots in intersectional inequities: B. Building empathy and the ability to deeply connect with local communities of interest; C. Developing leadership skills to nurture relationships with local communities, enhance multi-stakeholder partnerships, and manage diverse groups; D. Cultivating a growth mindset to manage and expect failures, learn over time, and develop resiliency; E. Practicing adaptability to rapidly iterate and respond to local contexts and feedback; F. Enhancing communication skills to effectively communicate with a broad range of communities, especially people with lived experience and potential partner; G. Strengthening entrepreneurial skills to raise funds for social innovation, develop innovative financing and sustainability approaches, and rapidly iterate ideas reported by participants attending the training workshop. Participants self-rated their competency in these seven main areas following workshop completion compared to their competency before training. The violin width represents the number of participants at a certain value. Solid lines indicate the median value, and dotted lines indicate the IQR. Statistical significance between groups was assessed using the Mann–Whitney U test.

**S8 Table:** Full Consensus Statements.

|  |
| --- |
| **Preamble** |
| 1. Social innovation can be defined as a community-engaged process that impacts health and social outcomes (Halpaap et al, Lancet GlobalHealth, 2020). Social innovation education and training can give local communities and health practitioners the key tools to solve problems relating to health. Currently limited evidence and consensus exists regarding core competencies that should be consistently achieved from social innovation in health training.<br>Although numerous education programmes exist globally, content and methods of delivery vary significantly, and most have been developed in the context of high-income countries.<br>The purpose of this survey is to identify social innovation learning competencies that can inform the delivery of teaching materials in social innovation in diverse settings, particularly resource constrained settings. The consensus statement which will be constructed from this project using modified Delphi methodology can be used by individuals learning about social innovation in health as well as research institutions, universities, and educators to build capacity for social innovation in health.<br>The primary target audience in this project is the individual learning about social innovation. We acknowledge that some competencies/mindsets/skills can be taught in a structured manner while others may be learnt from experiential learning or self-directed learning. Statements which refer to competencies that may be taught in structured social innovation training will be denoted with an * |
| **Mindsets** |
| 2. **Empathy** refers to the ability to deeply understand and share feeling that the target audience/user of any given innovation targets. While empathy may be difficult to teach in some circumstances, it is important for learners to have an awareness of its importance and focus on developing empathy so that the beneficiaries' needs are understood and met. |
| 3. **Resilience** refers to the ability to withstand difficulties and challenges. It refers to an ability to deal with failure and ambiguous circumstances. Developing resilience should be an important focus area for learners to develop in the context of social innovation in health and practical tools and guidance can be used to build resilience. |
| 4. **Adaptability** can ensure that social innovations are dynamic and thus can be implemented in different landscapes. Adaptability to rapidly iterate and respond to local contexts and needs is an important mindset to develop as part of social innovation training. |
| **Skills** |
| **Communication** |
| 5. **Pitching** is the ability to communicate verbally to gain support. Innovators who have better abilities to communicate their project, their needs and call for action assertively, have greater chances of success in seeking support. Pitching skills are therefore an important competency to become a competent social innovator. * |
| 6. **Community engagement** is the process of working collaboratively with groups of people whether they are connected by geographic proximity, special interest, or similar situations to address shared issues. By developing community partnership-building competencies, social innovation trainees will be better equipped to navigate the local context, align diverse interests, and mobilize the necessary resources and support to bring their ideas to life. Engaging communities is a key skill to develop in social innovation training programmes. * |
| 7. **Storytelling** is used to connect people to real life experiences. It has the power to connect people, convey real human experiences, and inspire others to action. Storytelling is an important skill which should be included in social innovation training content. * |
| **Knowledge** |
| **Intersectionality** |
| 8. Training programs should equip social innovators with the skills to **engage respectfully with different cultural norms, values, and experiences**. This includes developing cultural sensitivity, communication strategies, and the ability to facilitate cross-cultural collaboration. Lessons on navigating diverse cultural contexts are essential in social innovation training. * |
| 9. Understanding **health disparities** from an intersectional lens is crucial for social innovators to create innovations that can truly address the needs of the most vulnerable populations. Understanding health disparities related to age, ethnicity and other intersectional issues should be a core theme in social innovation training. * |
| 10. The **social determinants of health (SDH)** are the non-medical factors that influence health outcomes. They are the conditions in which people are born, grow, work, live, and age, and the wider set of forces and systems shaping the conditions of daily life. Equipping trainees with the skills to analyse and identify the specific social determinants at play in a given context will enable them to design more targeted, impactful solutions. This type of systems-level thinking, and root cause analysis is essential for social innovators to move beyond surface-level interventions and drive sustainable, systemic change. Skills to identify the social determinants that cause these health disparities are important to teach as part of social innovation training. * |
| 11. **Prioritizing indigenous talents** and wisdom sends a powerful message about the core values of social innovation which are community-centricity, co-creation, and a deep respect for local context and expertise. Training programs should emphasize the importance of humility, respect, and a willingness to learn from local community members. Social innovators must be equipped with the mindset and skills to identify, uplift, and collaborate with indigenous leaders, knowledge holders, and change-makers. Prioritizing indigenous talents, strength, and wisdom of local communities should be a core message in social innovation training. * |
| 12. Learning how to effectively **engage with community members, conduct needs assessments**, and gather insights about the key issues and pain points experienced by the target population is essential. Identifying needs/challenges of the community so that social innovations can be tailored to meet these needs is an important skill which should be learnt. * |
| **Research and Evaluation Skills** |
| 13. Teaching **evaluation of social innovation/impact assessment** including how to develop metrics and indicators that key stakeholders can understand, and value should be a component of training. * |
| 30. **Research skills including conducting literature reviews** are an important competency in social innovation in health. These skills include search strategies, accessing and interpretation of literature. * |
| 14. **Manuscript writing**, and publication (writing for scientific publication) can help to demonstrate legitimacy of social innovations in order to gain support. Manuscript writing is a key skill to learn to be a competent social innovator. * |
| 15. Effective media use (including social media) for information dissemination and includes skills like content creation, social media management, digital marketing, and data analytics. It should be an important learning objective in social innovation. |
| **Entrepreneurship** |
| 16. **Navigating funding pathways** such as grant writing, securing investment and business model knowledge is an important skill for social innovators to develop. * |
| 17. Lessons on **building and managing a team** may include content on topics like team formation, communication, conflict resolution, and performance management can equip social innovators with the competencies to build and lead high-functioning, interdisciplinary teams and should be part of social innovation in health learning content. * |
| 18. Sustainable innovations are solutions that can be maintained, replicated, and scaled to drive significant, durable change. Teaching on **creating sustainable innovation** that will be long-lasting is an important learning competency. * |
| 19. Lessons on **network management** can cover topics like maintaining effective communication, coordinating joint activities, and sustaining long-term collaborations. How to build and maintain networks/relationships is something that should be included in social innovation learning content. |
| **Participatory Methods** |
| 20. **Crowdsourcing** is an approach in which a group of individuals attempt to solve all or part of a problem, then implement exceptional solutions in the community. It leverages the collective knowledge, creativity, and problem-solving abilities of a diverse group of individuals to tackle complex challenges. Crowdsourcing to identify social innovations is a useful tool to teach. * |
| 27. **Community based participatory research** is reliant on building community partnerships and co-creation of solutions. Co-creation of solutions with the people who will be impacted can build trust, ownership, and buy-in from the target community. It can leverage existing community assets, resources and ensure accessibility, relevance, and long-term sustainability of innovations. By making community engagement a core part of the training process, social innovators can cultivate essential skills like stakeholder mapping, facilitation, conflict resolution, and collaborative problem-solving. Community based participatory research is an important approach to achieving social innovation in health competencies. * |
| **Ethics and Advocacy** |
| 21. Learning materials should provide in-depth coverage of relevant **ethical frameworks, guidelines, and best practices** to ensure social innovations in health are designed and implemented responsibly. This knowledge helps social innovators identify potential ethical risks, make well-reasoned decisions, and establish clear accountability measures. Integrating ethical considerations also fosters a deeper sense of social responsibility and commitment to the wellbeing of the communities served. Ethical considerations and pathways should be included in social innovation in health training. |
| 22. **Advocacy tools** may include stakeholder mapping, policy analysis, lobbying and campaigning, coalition building, and communication/messaging techniques. By cultivating advocacy competencies, social innovation training programs can help learners become more effective at influencing decision-makers, shifting mindsets, and mobilizing support for their health-focused initiatives. Advocacy tools should be taught to overcome barriers in local health systems to optimize success of social innovations. * |
| 29. Learning how to **navigate government policy including regulatory landscapes** is an important skill to be included in social innovation in health training. |

| Practical Tools |
| --- |
| 23. **User centred design** is the iterative design process in which designers focus on the users and their needs in each phase of the design process. Teaching user centred design is important to deliver comprehensive social innovation training* |
| 24. Although **mentoring** one-to-one can be difficult practically especially if resources are limited, it allows for tailored feedback, troubleshooting, and the cultivation of essential soft skills that are difficult to impart through classroom-based instruction alone. One-to-one or alternatively group mentoring is important to achieve core learning competencies. |
| 25. **Prototyping** allows social innovators to quickly test and iterate on their ideas, gather feedback, and refine their solutions before investing significant resources. Prototyping is a key skill to learn in social innovation training. * |
| 26. Using **generative play** i.e. playful activities and games to stimulate creativity and curiosity can be a useful tool to achieve important learning competencies in social innovation in heal |
| 28. **Definitions, theories, frameworks, case studies** are all key tools to achieve key social innovation competencies and skills. It is important to have resources that clarify what sets social innovation apart from other innovations by providing theoretical underpinnings. * |

